# 5G Radio-Frequency-Electromagnetic-Field Effects on the Human Sleep Electroencephalogram: A Randomized Controlled Study in *CACNA1C* Genotyped Healthy Volunteers

**DOI:** 10.1101/2024.12.16.24319082

**Authors:** Georgia Sousouri, Corinne Eicher, Rachele Maria D’ Angelo, Marie Billecocq, Thomas Fussinger, Mirjam Studler, Myles Capstick, Niels Kuster, Peter Achermann, Reto Huber, Hans-Peter Landolt

## Abstract

**Background:** The introduction of 5G technology as the latest standard in mobile telecommunications has raised concerns about its potential health effects. Prior studies of earlier generations of radiofrequency electromagnetic fields (RF-EMF) demonstrated narrowband spectral increases in the electroencephalographic (EEG) spindle frequency range (11-16 Hz) in non-rapid-eye-movement (NREM) sleep. However, the impact of 5G RF-EMF on sleep remains unexplored. Additionally, RF-EMF can activate L-type voltage-gated calcium channels (LTCC), which have been linked to sleep quality and EEG oscillatory activity.

**Objective:** This study investigates whether the allelic variant rs7304986 in the *CACNA1C* gene, encoding the α1C subunit of LTCC, modulates 5G RF-EMF effects on EEG spindle activity during NREM sleep.

**Methods:** Thirty-four healthy, matched participants, genotyped for rs7304986 (15 T/C and 19 T/T carriers), underwent a double-blind, sham-controlled study with standardized left-hemisphere exposure to two 5G RF-EMF signals (3.6 GHz and 700 MHz) for 30 min before sleep. Sleep spindle activity was analyzed using high-density EEG and the Fitting Oscillations & One Over f (FOOOF) algorithm.

**Results:** T/C carriers reported longer sleep latency compared to T/T carriers. A significant interaction between RF-EMF exposure and rs7304986 genotype was observed, with 3.6 GHz exposure in T/C carriers inducing a faster spindle center frequency in the central, parietal, and occipital cortex compared to sham.

**Conclusion:** These findings suggest 3.6 GHz 5G RF-EMF modulates spindle center frequency during NREM sleep in a *CACNA1C* genotype-dependent manner, implicating LTCC in the physiological response to RF-EMF and underscoring the need for further research into 5G effects on brain health.

## Introduction

Recent advances in mobile telecommunication established the fifth generation, “new radio” technology (5G) as the latest standard of world-wide wireless signal transmission technologies. The increasingly broad deployment of 5G raised widespread public concerns about its possible adverse health effects (Frey, 2021; Quoss et al., 2021). Particularly people who rate themselves as electromagnetic hypersensitive are concerned about sleep disturbances, headaches, and associated brain health impairments due to the non-ionizing radiation associated with radio-frequency electromagnetic field (RF-EMF) exposure (Baliatsas et al., 2014; Röösli et al., 2021). While the long-term health effects of EMF exposure are unclear, several independent studies demonstrated that 2G-4G EMF acutely alter electroencephalographic (EEG) ∼ 9-16 Hz oscillations in wakefulness and sleep (Borbély et al., 1999; Croft et al., 2008; Huber et al., 2000; Loughran et al., 2005, 2019; Regel et al., 2007; Schmid, Loughran, et al., 2012; Schmid, Murbach, et al., 2012). More specifically, enhanced spindle or “sigma” activity (∼11-16 Hz) in non-rapid-eye-movement (NREM) sleep is among the most consistent effects of 2G-4G RF-EMFs. Nevertheless, there are striking inter-individual differences in how volunteers respond to acute EMF exposure and some EMF studies failed to observe significant effects on the sleep EEG (Bosch-Capblanch et al., 2024; Danker-Hopfe et al., 2020; Hinrikus et al., 2022; Loughran et al., 2012; Lustenberger et al., 2015; Schmid, Loughran, et al., 2012). Pulse modulation of 2G-4G EMF signals appears to be critical for induction of sleep EEG alterations (Huber et al., 2002; Regel, Tinguely, et al., 2007).

The 5G technology operates on higher carrier frequencies (frequency range 1: 450-6000 MHz), wider bandwidths (up to 120 MHz), and more complex signal characteristics than the previous technologies. The wider bandwidths may mimic continuous wave exposure yet distinct features for low-latency and low-power communication may introduce short, pulsed waveforms. Thus, it is unknown whether 5G EMFs affect the sleep EEG.

The high inter-individual variability and high intra-individual stability of the sleep EEG changes induced by 2G-4G EMF hints to a possible genetic predisposition that may provide an avenue to elucidate their biological underpinnings. What could be a biological mechanism, by which EMFs affect human brain physiology? While EMF exposure does not provide sufficient energy to break chemical bonds, ample evidence demonstrates that EMF can alter cellular properties in many organs of the body. Accordingly, EMF exposure depolarizes the membrane potential of excitable cells in skeletal muscles, heart, brain, and endocrine organs, which activates voltage-gated calcium ion (Ca^2+^) channels and leads to increased intra-cellular Ca^2+^ concentration (Panagopoulos et al., 2002; Kim et al., 2019; Pall, 2013; Piacentini et al., 2008; Rao et al., 2008). The Ca^2+^ influx drives processes such as hormone secretion, neurotransmitter release, muscle constriction, gene transcription, and neuronal activity (Dolphin, 2016; Neher & Sakaba, 2008). Thus, these ion channels are of critical importance for virtually all brain functions, and their dysfunction can give rise to various vascular and brain diseases (Simms & Zamponi, 2014; Zamponi, 2016). Interestingly, distinct allelic variants in the third intron of the gene *CACNA1C* encoding the α1C subunit of the L-Type, voltage-gated calcium channel (LTCC), associated with prolonged self-rated sleep latency and reduced sleep quality (Byrne et al., 2013; Kantojärvi et al., 2017; Parsons et al., 2013). This subunit determines the voltage sensitivity and conductance of LTCC which are expressed on almost all types of neurons in the brain and regulate neuronal firing, learning and memory, addictive behaviors, and neuronal development (Dolphin, 2016; Striessnig et al., 2014; Zamponi et al., 2015). In addition, the LTCCs are known molecular substrates of sleep EEG oscillations in vitro and in vivo (Kumar et al., 2015).

The variations in exposure parameters — such as frequency, duration, intensity, and the specific characteristics of the signal source — of different generations of RF-EMF complicate the process of establishing a consistent and reliable understanding of how RF-EMF affects human electrophysiology. Another factor possibly contributing to some inconsistent results in EMF effects on the human sleep EEG may be the limitations of the pre-defined, fixed-band quantification of oscillatory EEG activity. The common averaging of power spectra ignores inherent individual variability in the oscillatory characteristics, which may mask or eliminate otherwise observable effects. For the topographical characterization of neural oscillations, analysis of pre-defined spectral bands is highly problematic. For example, sleep spindles exhibit a distinct spatial pattern, with slower spindles prevailing in frontal cortical areas and faster spindles in the centro-occipital areas (Fernandez & Lüthi, 2020; Werth et al., 1997). Hence, analysis of pre-defined frequency bands may result in misleading estimations of power from the area under the curve. A recently published methodological approach aims to tackle this problem, the Fitting Oscillations & One Over f (FOOOF) analysis (Donoghue et al., 2020). The FOOOF algorithm enables the separate quantification of broadband, aperiodic background power and distinct, periodic, oscillatory components of neural power spectra. The FOOOF analysis provides a validated, intuitive method for reliable and informative extraction of the individual spectral EEG characteristics. More specifically, the NREM sleep EEG power spectrum displays a characteristic, decreasing trend across increasing frequencies (1/f like) with a distinctive ‘peak’ of power in the frequency range of sleep spindles. The FOOOF algorithm fits Gaussian models to parameterize the power spectra, allowing for the extraction and characterization of various components of the “spindle peak.”

Based on this background, the goal of this study was to investigate whether pre-sleep exposure to realistic, standardized 5G EMF signals affects the spectral characteristics of spindles in the NREM sleep EEG and whether the EMF-induced changes are modulated by the variant rs7304986 of the *CACNA1C* gene.

## Materials and methods

### Participants

Thirty-four healthy, right-handed volunteers were enrolled in this study. Based on specific inclusion and exclusion criteria, they were selected among the 2,040 participants of an observational study on self-rated electrohypersensitivity and sleep (Eicher et al., 2024). As described in detail, all participants returned a saliva collection kit (OG-500 by DNA Genotek Inc.) for DNA extraction and genotyping of allelic variants rs7304986 and rs16929277 of *CACNA1C* (Eicher et al., 2024). For this study, we identified the T/C allele carriers of rs7304986 and prospectively matched them based on sex, age and body-mass-index with a corresponding T/T allele carrier. Because of dropouts, the final study groups consisted of 15 T/C and 19 T/T allele carriers of the rs7304986 variant. All participants completed a series of questionnaires addressing their mobile phone usage, medication intake, sleep patterns, general and neurological health. Additionally, we employed the Pittsburgh Sleep Quality Index (PSQI) (Buysse et al., 1989) to assess subjective sleep quality, the Epworth Sleepiness Scale (ESS) (Johns, 1991) to estimate daytime sleepiness, and the Munich Chronotype Questionnaire (MCTQ) (Allebrandt & Roenneberg, 2008) to quantify diurnal preference. The subjective sensitivity to EMF was assessed using a questionnaire developed by (Röösli et al., 2010). Those participants who endorsed the question “Are you electro-hypersensitive?” were classified as “EHS”. Those who negated this question but confirmed that they “think to develop detrimental health symptoms due to electromagnetic pollution in everyday life” were classified as “attributers”. All other participants were categorized as “non-EHS”.

### Experimental Protocol

All study procedures were approved by the Cantonal Ethics Committee (BASEC-ID: 2016- 02049) and conducted in accordance with the Declaration of Helsinki. The study protocol was registered on the Swiss National Clinical Trials Portal (# SNCTP000002285) and ClinicalTrials.gov (# NCT03074617). Written informed consent was obtained from all participants prior to participation. Prior to the study, all participants adhered to behavioral instructions aimed at minimizing external influences on sleep. For three days prior to each experimental session, they abstained from alcohol and caffeine and maintained consistent bedtimes from 11:00 PM to 7:00 AM, with compliance verified by daily sleep diaries and wrist actigraphy monitoring (Actiwatch Type AWL, CamNtech, Cambridge, UK). The experimental nights of the female participants took place in the first three weeks of their menstrual cycle, to ensure that hormonal fluctuations would not confound the results of the study. Mobile phone use was restricted from the evening before the experimental night, and all participants refrained from sports or sauna visits on the days of experimental nights. Throughout the study, participants did not travel across more than two time zones, engage in night shift work, take medications, smoke cigarettes, use drugs, or participate in other clinical studies.

All participants completed three experimental nights with different, standardized exposure conditions according to a randomized, double-blind, cross-over design: 1) 30-min, pre-sleep exposure to an active 5G EMF at a carrier frequency of 700 MHz, 20 MHz bandwidth, and 12.5 Hz applied power control, 2) 30-min, pre-sleep exposure to an active 5G EMF at a carrier frequency of 3.6 GHz, 100 MHz bandwidth, and 12.5 Hz applied power control, and 3) a 30- min sham exposure without an active field. Exposure started 60 min before bedtime and was applied to the left cerebral hemisphere of the participants (not known to the participants). Each experimental night was separated by one week, and the sequence of conditions was counterbalanced among the subjects. During exposure, the participant was seated on a chair and the participant’s head was positioned between two boxes. Two patch antennas were inside the box on the left side of the head, centered 42 mm vertically above the auditory canal and 115 mm distance to the left side of the head, targeting the electromagnetic field primarily at the left hemisphere. Directly after the exposure, the high-density (hd)-EEG net was hooked up on the participants’ head strictly observing a 30-min time window between the end of the exposure and the start of the sleep recording (lights out). During the preparation of the participants, a subjective post-exposure survey was administered, to assess whether they perceived a field or not, and whether they had complaints to report during or immediately after the exposure had occurred.

### 5G Signal Characterization

All exposure conditions were administered with the same exposure system (sXh5G), provided by the IT’IS Foundation for Research on Information Technologies in Society (IT’IS Foundation, Zurich, Switzerland), which ensures controlled and well-characterized 5G EMF exposure. Following detailed simulated dosimetry (Figure 1), the signal intensity was calibrated to ensure the specific absorption rate (SAR) for the head (averaged over 10 g of tissue) did not exceed 2 W/kg. The active field remained within the SAR limit for the general population established by the International Commission on Non-Ionizing Radiation Protection (ICNIRP) and posed no known health risks. The two active fields administered are 5G uplink signals generated in the 5G frequency range. The lower frequency signal has a carrier frequency of 700 MHz, 20 MHz bandwidth, Frequency Division Duplexing/Orthogonal Frequency-Division Multiplexing (FDD/OFDM) with 24 resource blocks, 16 time slots, 60 kHz sub-carrier spacing and Quadrature Phase Shift Keying (QPSK) modulation, with an output power of 4.28 W. The higher frequency signal has a carrier frequency of 3.6 GHz, 100 MHz bandwidth, Time Division Duplexing (TDD)/QPSK OFDM with 135 resource blocks, 16 time slots, 60 kHz sub-carrier spacing and QPSK modulation, with an output power of 1.63 W. In the signals used, only uplink communication is implemented, and all frames are identical with 16 time slots. Both signals have identical power control applied that introduces low frequency amplitude modulation at 12.5 Hz on top of the modulation due to the occupied time slots which have a dominant power modulation frequency of 200 Hz resulting in a 14.2 dB peak to average power ratio (PAPR) (Supp. Figure 1, 2 & 3). The exposure levels in grey and white matter, thalamus and all tissues in the brain averaged over 0.125 g which is a cube of side length ∼5 mm are reported in Table 1 for both 700 MHz and 3.6 GHz. Peak exposures are in the cortical tissue closest to the antenna, the most noticeable feature being the much higher rate of decay of the SAR at the higher frequency. During sham exposure no signal is emitted from the antenna. The system has been extensively tested and validated through numerical and experimental dosimetry of the antenna, including uncertainty and variability analyses (Supp. Table 1, 2, 3 & 4), measurement of psSAR10g in a twinSAM phantom, measurement of the electric field in free space, and quantitative comparison of the validation measurement and the numerical dosimetry. Hence, the exposure in the experiments can be compared with the actual exposure of users in real networks. To exclude the possibility of any interference with other sources of electromagnetic radiation, the shielding of the sleep laboratory was confirmed by validated measurements conducted by the IT’IS Foundation. All WiFi access points in the surrounding floors of the building were deactivated and all mobile phones, smart watches, tablets, trackers and laptops in the vicinity of the sleep laboratory were turned off.

**Figure 1.**
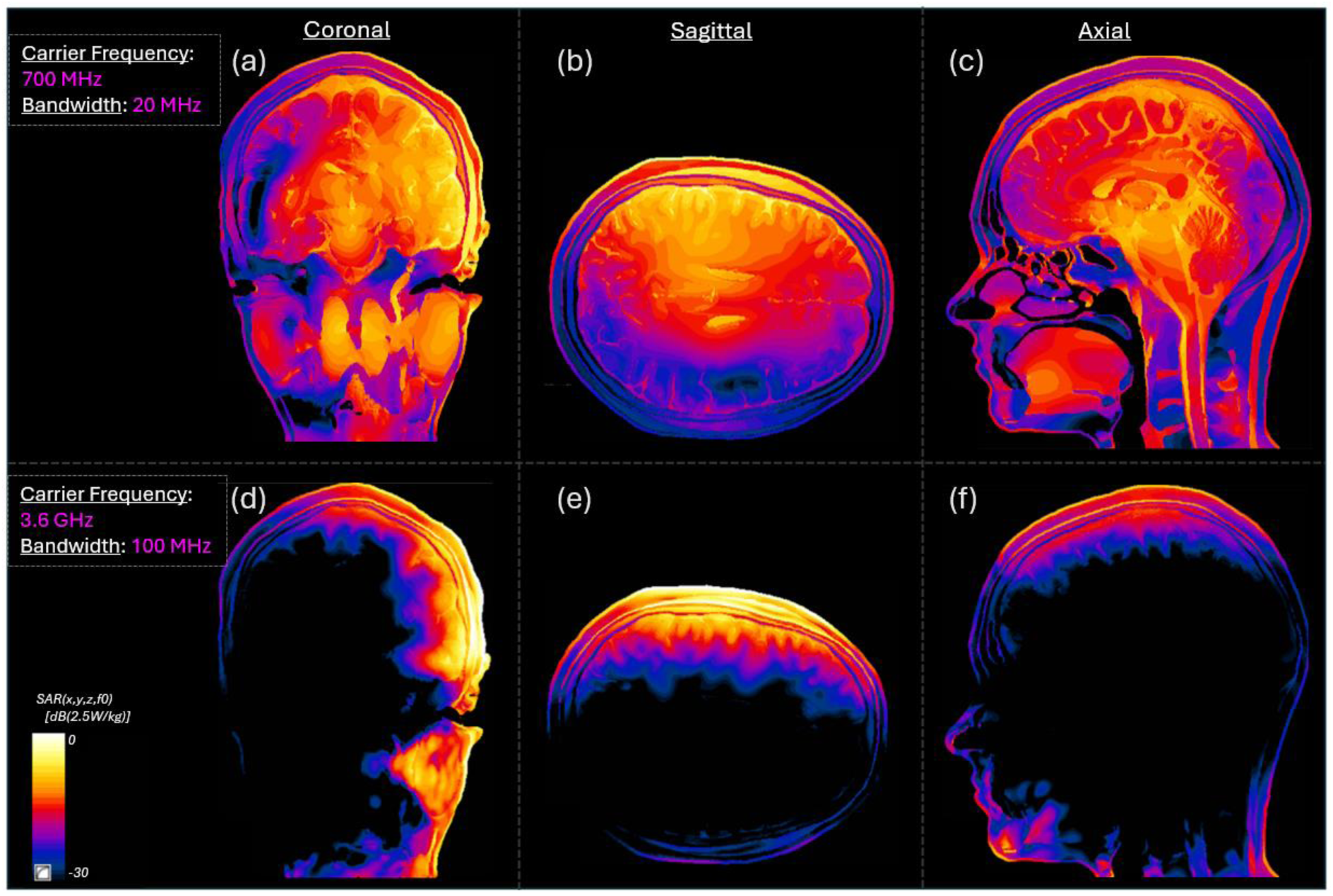
Simulated SAR Value Distributions for Both Active Fields. Top row: simulated Specific Absorption Rate (SAR) cross-sections at 700 MHz and 1 Watt conducted input power. (a) coronal, (b) axial, (c) sagittal view. Bottom row: simulated SAR cross-sections at 3.6 GHz and 1 Watt conducted input power. (d) coronal, (e) axial, (f) sagittal view. The color bar (30 colors) indicates the SAR in dB scale −30 to 0 dB with reference to 2.5 W/kg.

**Table 1:** Peak spatial SAR normalized for 2-W maximal psSAR10g.

| Tissue(s) | psSAR 0.125g W/kg |  |
| --- | --- | --- |
|  | 700 MHz | 3.6 GHz |
| Grey Matter | 2.55 | 1.23 |
| White Matter | 1.68 | 0.49 |
| White and Grey Matter | 2.48 | 1.19 |
| Whole Brain | 2.48 | 1.19 |
| Thalamus | 0.91 | 0.004 |

*Simulated Specific Absorption Rate (SAR) values averaged over 0.125 g, equivalent to cubes of 5mm side length, for regions of interest in the head, for both the 700 MHz and the 3.6 GHz signals. Values are normalized to the maximal peak spatial SAR averaged over 10g of tissue of 2 W as used in the experiments.*

### High-density Sleep Electroencephalography

During the experimental sessions, we employed 128-channel Electrical Geodesics Sensor Nets for all-night hd-EEG recordings (Electrical Geodesics Inc., EGI, Eugene, OR). To allow for parallel recordings of two participants in each experimental night, we used two types of amplifiers of different generations (Net Amps 300 and 400 series). We combined the hd-EEG recordings with electrooculographic (EOG) and submental electromyographic (EMG) recordings for visual scoring of sleep states. We adjusted the net to the vertex and the mastoids and filled all electrodes with an electrolyte gel (ECI Electro-Gel, Electro-Cap International, Inc., Eaton, OH) to ensure high conductance and maintain good signal quality throughout the night. Impedances of all EEG electrodes were below 50 kΩ at the start of the recording and were checked again in the morning. We referenced all channels to Cz and the sampling rate was set at 500 Hz. To avoid potential systematic differences between the two recording systems used and to ensure consistency and reliability of the results, we retrospectively re-calibrated all EEG recordings before data analyses.

### Sleep EEG scoring, artifact removal and cycle analysis

For the sleep scoring we applied the following pre-processing: using the default Hamming-windowed FIR filter implemented in EEGLAB 2021.0 (function pop_eegfiltnew), the raw EEG and EOG data were low-pass filtered at 36 Hz and high-pass filtered at 0.3 Hz. Using the same filter function, the EMG was low-pass filtered at 100 Hz and high-pass filtered at 10 Hz. We down-sampled the filtered EEG, EOG and EMG data to 150 Hz. Using ‘Visbrain’ software (Combrisson et al., 2019) a sleep expert visually scored sleep states in 30-s epochs according to the American Academy of Sleep Medicine (AASM) criteria (Berry et al., 2017), with subsequent verification by another sleep expert (both blind to the experimental conditions). For the artifact removal, we applied the following pre-processing: the raw sleep EEG was conditioned with a notch-filter at 50 H z (Finite Impulse Response (FIR) filter, kaiser window, zero-phase, filter order 560, cut-off frequencies (−6 dB) at 47 and 53 Hz) to attenuate the power line noise and its harmonics. The NREM EEG was low-pass filtered at 40 Hz using the default FIR filter implemented in EEGLAB v2021.1 (Hamming window, zero-phase, filter order 166, cut-off frequency (–6 dB) at 45 Hz), and high-pass filtered at 0.9 Hz (FIR filter, kaiser window, zero-phase, filter order 4978, cut-off frequency (–6 dB) at 0.6 Hz). Data was re-referenced to the average of the mastoids and down-sampled to 128 Hz. We utilized ‘High-Density-SleepCleaner’, a semi-automatic procedure (Leach et al., 2023) to efficiently detect and exclude artifactual epochs and EEG channels in the NREM sleep EEG. This is a Graphical User Interface (GUI) where outlier values are visually identified by the user based on four Sleep Quality Markers (SQMs): 1) delta power (0.5-4.5 Hz; from robustly z-standardized EEG data), 2) beta power (20-30 Hz; from robustly z-standardized EEG data), 3) the maximum squared deviation in amplitude from the mean EEG signal, and 4) delta power (from raw, not robustly z-standardized, EEG data). All SQMs were computed from both the original reference EEG data (Cz) and the average-referenced EEG data (Leach et al., 2023). Finally, artefactual channels were recovered by nearby channels using epoch-wise spherical interpolation (EEGLAB 2021.0). Following the sleep staging and the cleaning of the sleep epochs, sleep cycle analysis was performed using the ‘Sleep Cycles’ package in R (Blume & Cajochen, 2021) which is based on criteria originally proposed by (Feinberg & Floyd, 1979). Accordingly, NREM sleep periods have had a minimal duration of 15 min, which can include wakefulness and up to 5 min of REM sleep. Except for the first REM sleep period where no minimum duration criterion was applied, any REM sleep period must be at least 5 min long (Feinberg & Floyd, 1979). Due to technical issues with the EEG amplifier, the last part of the night was compromised in some recordings: T/T carriers in the 3.6 GHz exposure (n = 2), T/C carriers in the sham (n = 1), 700 MHz (n = 3), and 3.6 GHz exposure (n = 1). These recordings were excluded from whole-night analysis; however, they were retained in the analysis of the first sleep cycle.

### Sleep EEG spectral analysis

For our main analysis, the pre-processed sleep EEG data following artifact removal was re-referenced to the average of the mastoids. All signals were spectrally decomposed by computing power spectral density (PSD) values for each epoch using the Welch method (pwelch function) in Matlab (R2021b, The Math Works Inc., Natick, MA, USA). For each 30-s epoch, 4-s Hanning windows were used with a 50% overlap resulting in a frequency resolution of 0.25 Hz. Channels from the outer ring of the electrode array that do not capture EEG activity were excluded from the analysis. Analysis was conducted on a subset of EEG channels deemed relevant for the study of brain activity, totaling 109 channels.

### Fitting Oscillations & One Over f (FOOOF) analysis

To comprehensively disentangle the effects of 5G exposure on the sleep EEG, we used the Fitting Oscillations & One Over f (FOOOF) toolbox (Donoghue et al., 2020). The FOOOF algorithm parameterizes the neural power spectra into their aperiodic and periodic/oscillatory components by fitting gaussian distributions through an iterative process (Supp. Figure 4). For each participant, each night recording, and each channel, the mean NREM (combined N2 and N3) sleep EEG power spectra of the whole night and separately for each sleep cycle were fit into the algorithm for the detection of sleep spindle peaks. The FOOOF model was parameterized as follows: peak width limits 1-5 Hz (to span the sigma frequency range of 11- 16 Hz), maximum number of detected peaks was set to 4, and the minimum peak height to 0.05 log(power). Peak detection was performed in the frequency range of 1-30 Hz. For the spindle peak detection in the NREM sleep EEG, the following properties were extracted and stored for further analyses: peak center frequency [Hz], relative peak power [log_10_(μV^2^/Hz)], and peak bandwidth [Hz]. Peaks of interest were selected based on frequency range and power criteria. More specifically, all the peaks with a center frequency in the sigma range (11- 16 Hz) were identified. If more than one peak was detected, then the one with the highest relative power was selected. This criterion helped to identify the most prominent spindle peak in the power spectrum and avoid false positives in the peak detection.

### Statistics

We used linear mixed models (LMM) with nested data structures, to describe the data in our repeated measures design (lme function from nlme R package). We applied Wald Chi-squared test statistics for the description of goodness of fit of the LMM. For the LMM, the participant was treated as the random variable, varying in the intercept and nested within the participant matching. The fixed effects were ‘genotype’ (T/T and T/C genotypes), ‘exposure’ (conditions S0 [sham], E7 [700 MHz carrier frequency, 20 MHz bandwidth] and E3 [3.6 GHz carrier frequency, 100 MHz bandwidth]), and ‘night’ (experimental nights 1, 2, and 3). We used the maximum likelihood method for the estimation of the parameters of interest. In case of more than one variable, we performed bi-directional stepwise regression using the Akaike Information Criterion (AIC) for model comparison (stepAIC function in MASS R package). The stepwise principle for model construction is an automatic procedure of selecting the regression model that best fits the data. Initially, all the candidate predictive variables were included in the model, and then the bi-directional stepwise regression was applied, to estimate the final model that included the variables and/or their interactions that best explained the variation of the outcome. In short, an initial model was defined only by the intercept. Next, the predictor that best predicted the outcome was selected according to the highest correlation with the outcome. If this predictor improved the ability of the model to predict the outcome, then it was retained in the model. Each time a predictor was added, a removal test was computed of the least useful predictor. Then, a second predictor is selected by using semi-partial correlations with the outcome as a criterion. At each step, the resulting models were compared to each other, using the AIC which was computed as AIC = –2(log-likelihood) + 2k, where k was the number of model parameters including the intercept and the log-likelihood was a measure of model fit. The lower the AIC, the better the fit of the model. The advantage of this method is that it provides an objective way to estimate the LMM. We confirmed normality using Shapiro-Wilk tests and used two-tailed, paired t-tests for post-hoc comparisons. In cases of multiple testing, we used the False Discovery Rate (FDR) Benjamini & Hochberg correction. We provide descriptive values of mean and standard error of the mean (± SEM; unless reported otherwise) and 95% confidence intervals (CI). We performed all statistical analyses in RStudio, R-4.0.2.

For the high-density EEG topographical comparisons, nonparametric, cluster-based permutation statistics were used, applying a suprathreshold cluster analysis to control for multiple comparisons (Nichols & Holmes, 2003). Briefly, for each topographical statistical comparison, the condition label was randomly permuted between the contrasting groups, and paired, Student’s t-tests were performed. For each permutation, the maximum size of resulting clusters of neighboring electrodes reaching a t-value above a critical value was computed to form a cluster size distribution. From this cluster size distribution, the 97.5th percentile was defined as the critical cluster size threshold. Only electrodes reaching a t-value beyond the critical value (CV) and located within a cluster larger than the critical cluster size threshold were considered significant (paired Student’s t-tests; T/T: CV = 2.101, corresponding to a = 0.05 for the given degrees of freedom, number of permutations 1000, n = 19; T/C: CV = 2.145, corresponding to a = 0.05 for the given degrees of freedom, number of permutations 1000, n = 15). We report Cohen’s d effect sizes. Permutation statistics for the topographical comparisons were performed in Matlab (R2021b, The Math Works Inc., Natick, MA, USA).

## Results

### Demographics of study sample

The major demographic characteristics of the two genotype groups are summarized in Table 2. Both groups reported good sleep quality (mean PSQI score < 5), normal daytime sleepiness (mean ESS score < 10), and no extreme chronotype. A minority in both genotype groups considered themselves as being electro-hypersensitive. Interestingly, corroborating previous findings (Byrne et al., 2013), the T/C allele carriers of *CACNA1C* reported a longer subjective latency to fall asleep than the T/T allele carriers.

**Table 2:** Demographic characteristics of the study participants.

|  | **T/T carriers** | **T/C carriers** | **p-value** |
| --- | --- | --- | --- |
| Sample size (n) | 19 | 15 | - |
| Sex ratio (female/male) | 16/3 | 14/1 | - |
| Age (years) | 24.7 ± 2.2 | 25.2 ± 2.7 | 0.56 |
| Body-mass-index (kg/m<sup>2</sup>) | 22.7 ± 1.7 | 22.1 ± 2.7 | 0.43 |
| Sleep quality (PSQI) | 3.3 ± 1.2 | 3.8 ± 1.9 | 0.31 |
| Daytime sleepiness (ESS) | 5.8 ± 2.2 | 5.6 ± 2.3 | 0.75 |
| Diurnal preference (MCTQ; hh:mm) | 03:49 ± 00:46 | 03:42 ± 01:11 | 0.71 |
| Preferred morning/evening types (%) | 53/47 | 58/42 | 0.33 |
| Habitual bedtime (hh:mm) | 23:05 ± 0:43 | 22:44 ± 0:53 | 0.20 |
| Habitual rise time (hh:mm) | 07:23 ± 0:59 | 07:10 ± 1:25 | 0.62 |
| Reported time to fall asleep (min) | 18 ± 9 | 29 ± 20 | **0.04** |
| Reported sleep duration (h:mm) | 7:49 ± 35 | 7:31 ± 48 | 0.21 |
| Electro-hypersensitivity (EHS) | Yes: 10.5%<br>No: 47.4%<br>Attributers: 42.1% | Yes: 6.7%<br>No: 60.0%<br>Attributers: 33.3% | 0.44 |

*Values represent means ± standard deviations or percentages per genotype group. T/T and T/C carriers: alleles of genetic variant rs7304986 of the gene CACNA1C. PSQI = Pittsburgh Sleep Quality Index; ESS = Epworth Sleepiness Scale; MCTQ = Munich ChronoType Questionnaire; EHS = Perception of electro-hypersensitivity according to Rösli et al. (2010). If the mid-sleep time on the MCTQ is earlier than 04:00, the participant is considered as preferential morning type, otherwise as preferential evening type. The p-values refer to two-tailed paired t-tests.*

### Sleep Variables

The statistical analysis of all-night sleep variables revealed an interaction between ‘exposure’ and ‘genotype’ (χ^2^(Df) = 7.2(2), Pr(>χ^2^) = 0.027; Df = degrees of freedom) for the combined N2 and N3 sleep stages (Table 3). The interaction factor was not included in the models for the rest of the sleep variables. In line with the anticipated first-night effect commonly observed in sleep laboratory settings (Le Bon et al., 2001), the factor ‘night’ showed a significant effect in the LMM models (Supp. Table 5). Nevertheless, post-hoc t-tests did not indicate any significant differences between the groups and/or the experimental conditions.

**Table 3:**
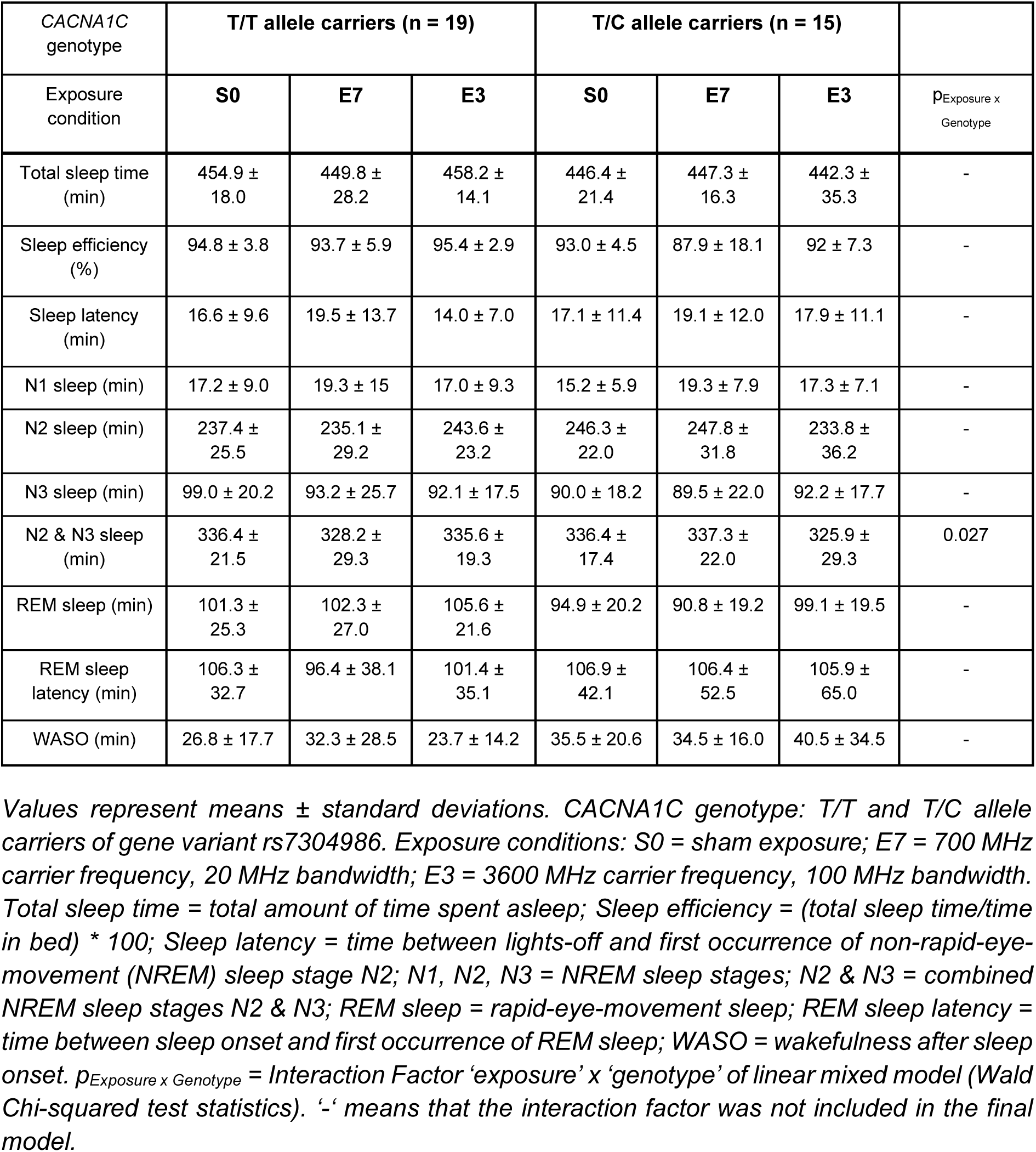
All-night sleep parameters for the three experimental conditions of both genotypes.

| CACNA1C genotype | T/T allele carriers (n = 19) |  |  | T/C allele carriers (n = 15) |  |  |  |
| --- | --- | --- | --- | --- | --- | --- | --- |
| Exposure condition | S0 | E7 | E3 | S0 | E7 | E3 | p<sub>Exposure x Genotype</sub> |
| Total sleep time (min) | 454.9 ± 18.0 | 449.8 ± 28.2 | 458.2 ± 14.1 | 446.4 ± 21.4 | 447.3 ± 16.3 | 442.3 ± 35.3 | - |
| Sleep efficiency (%) | 94.8 ± 3.8 | 93.7 ± 5.9 | 95.4 ± 2.9 | 93.0 ± 4.5 | 87.9 ± 18.1 | 92 ± 7.3 | - |
| Sleep latency (min) | 16.6 ± 9.6 | 19.5 ± 13.7 | 14.0 ± 7.0 | 17.1 ± 11.4 | 19.1 ± 12.0 | 17.9 ± 11.1 | - |
| N1 sleep (min) | 17.2 ± 9.0 | 19.3 ± 15 | 17.0 ± 9.3 | 15.2 ± 5.9 | 19.3 ± 7.9 | 17.3 ± 7.1 | - |
| N2 sleep (min) | 237.4 ± 25.5 | 235.1 ± 29.2 | 243.6 ± 23.2 | 246.3 ± 22.0 | 247.8 ± 31.8 | 233.8 ± 36.2 | - |
| N3 sleep (min) | 99.0 ± 20.2 | 93.2 ± 25.7 | 92.1 ± 17.5 | 90.0 ± 18.2 | 89.5 ± 22.0 | 92.2 ± 17.7 | - |
| N2 & N3 sleep (min) | 336.4 ± 21.5 | 328.2 ± 29.3 | 335.6 ± 19.3 | 336.4 ± 17.4 | 337.3 ± 22.0 | 325.9 ± 29.3 | 0.027 |
| REM sleep (min) | 101.3 ± 25.3 | 102.3 ± 27.0 | 105.6 ± 21.6 | 94.9 ± 20.2 | 90.8 ± 19.2 | 99.1 ± 19.5 | - |
| REM sleep latency (min) | 106.3 ± 32.7 | 96.4 ± 38.1 | 101.4 ± 35.1 | 106.9 ± 42.1 | 106.4 ± 52.5 | 105.9 ± 65.0 | - |
| WASO (min) | 26.8 ± 17.7 | 32.3 ± 28.5 | 23.7 ± 14.2 | 35.5 ± 20.6 | 34.5 ± 16.0 | 40.5 ± 34.5 | - |

Values represent means ± standard deviations. CACNA1C genotype: T/T and T/C allele carriers of gene variant rs7304986. Exposure conditions: S0 = sham exposure; E7 = 700 MHz carrier frequency, 20 MHz bandwidth; E3 = 3600 MHz carrier frequency, 100 MHz bandwidth. Total sleep time = total amount of time spent asleep; Sleep efficiency = (total sleep time/time in bed) \* 100; Sleep latency = time between lights-off and first occurrence of non-rapid-eye-movement (NREM) sleep stage N2; N1, N2, N3 = NREM sleep stages; N2 & N3 = combined NREM sleep stages N2 & N3; REM sleep = rapid-eye-movement sleep; REM sleep latency = time between sleep onset and first occurrence of REM sleep; WASO = wakefulness after sleep onset. $p_{\text{Exposure} \times \text{Genotype}}$ = Interaction Factor 'exposure' x 'genotype' of linear mixed model (Wald Chi-squared test statistics). '-' means that the interaction factor was not included in the final model.

### Topographical validation of sleep spindle peak parameterization

Initially, we confirmed the validated sleep spindle characterization of the FOOOF algorithm. Topographical maps of the derived spindle peak parameters in N2 and N3 all-night, sleep-EEG neural power spectra in the sham condition in all participants are depicted in Figure 2. The left panel (Figure 2A) shows that the center oscillatory frequency of sleep spindles is higher in central and occipital cortical regions (∼13-16 Hz) while frontal spindles oscillate in lower frequencies (∼11-13 Hz). The middle panel (Figure 2B) highlights that adjusted sigma power of the spindle peak relative to the 1/f like background spectral trend is highest in central, centro-occipital regions with spindle power being reduced towards the frontal cortical areas. The right panel (Figure 2C) illustrates that the bandwidth of sleep spindles is wider in frontal areas while spindle oscillatory frequency range becomes narrower in central and occipital cortical regions. These maps confirm the conventional topographical characterization of sleep spindles (Fernandez & Lüthi, 2020) and validate the usefulness of the FOOOF algorithm to derive sleep spindle characteristics from the NREM sleep EEG.

**Figure 2.**
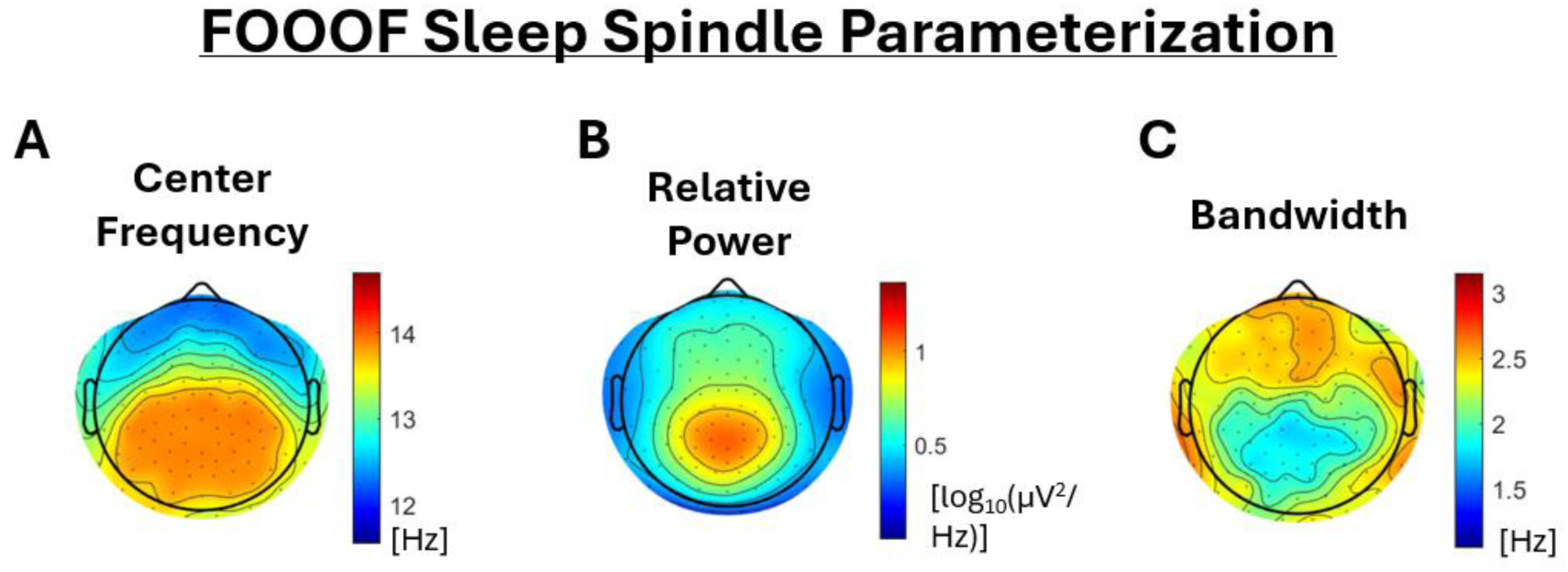
Sleep Spindle Peak Parameterization. Topographical characterization of FOOOF spindle peak detection (n = 33; sham condition). Left panel: center frequency of the detected spindle peaks (Hz). Middle panel: spindle peak power relative to the 1/f background activity (log_10_(μV^2^/Hz)). Right panel: bandwidth of detected spindle peaks (Hz). Different color-codes refer to each panel.

### Single-channel, all-night EEG

For a first visual inspection of the EEG power changes, we plotted the all-night, broadband (1- 25 Hz, 0.25-Hz bins) EEG power density spectra of the channel C3 referenced to the right mastoid (C3-RM), ipsilateral to the exposure site, in NREM sleep after the 30-min pre-sleep 5G RF-EMF exposure as the ratio to the power spectrum after sham exposure. In the T/C allele carriers of *CACNA1C* and after 3.6 GHz exposure, we observed a maximum increase in power spectral density of 23.9 ± 8.45 % (95% CI: [5.8 %, 42.1 %]) at 14.75 Hz following a maximum decrease of −15.9 ± 4.6 % (95% CI: [-6 %, −25.7 %]) at 13 Hz, which is evident as a profound spike in the ratio plot ‘TC-3.6 GHz/Sham’ (Figure 3A, bottom right panel). Note that a smaller spike-like trace is also evident in the ratio plot ‘TC-700 MHz/Sham’ (Figure 3A, bottom left panel) in the same spindle frequency range with a maximum power increase of 10 ± 7.4 % (95% CI: [-5.9 %, 25.8 %]) at 14.5 Hz and a decrease of −6.1 ± 3.9 % (95% CI: [-14.5 %, 2.3 %]) at 12.75 Hz. Statistical analysis did not show any significant difference from sham in any frequency bin.

**Figure 3.**
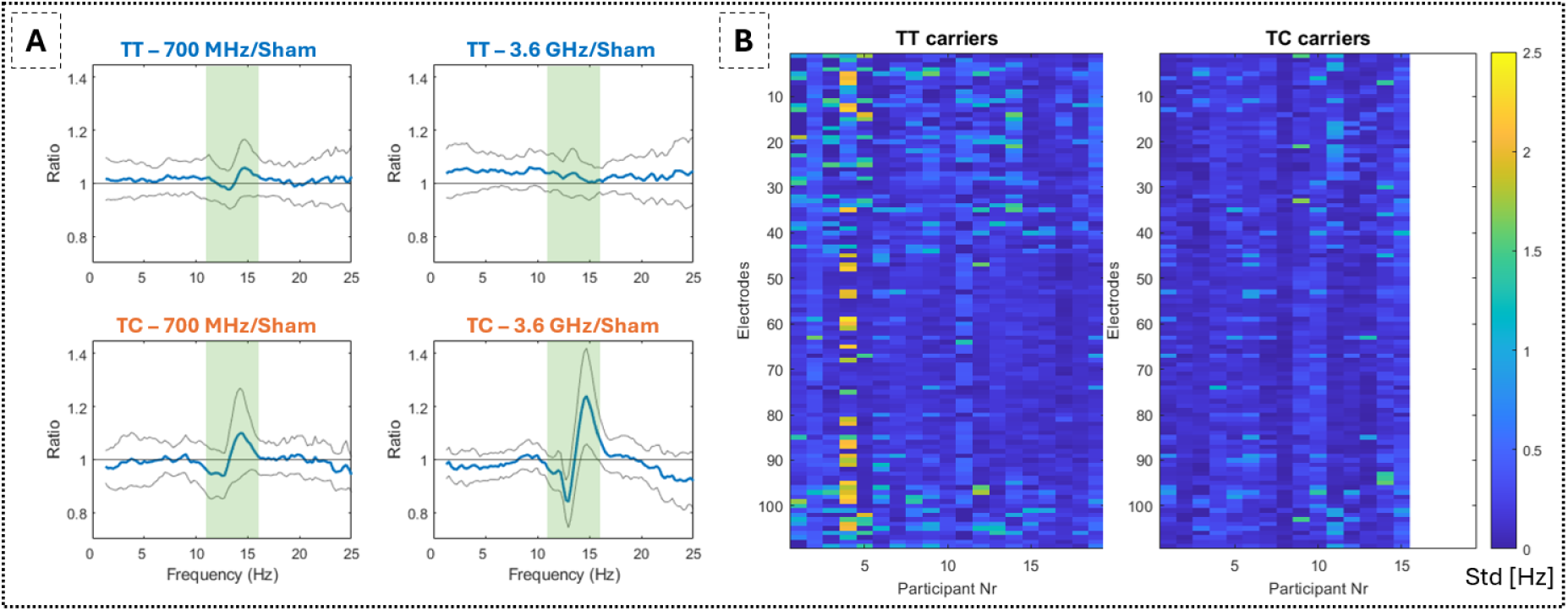
A) Mean ratio (blue lines) of whole-night, sleep EEG power density spectra (1-25 Hz, 0.25-Hz bins) between exposure conditions (700 MHz and 3.6 GHz) and sham in T/T (n = 19) and T/C (n = 15) allele carriers of *CACNA1C* in EEG derivation C3 referenced to the right mastoid (C3-RM). Green shading indicates the spindle frequency range (11-16 Hz). Grey lines indicate the 95% CI. B) Intra-subject standard deviation of the selected spindle peak frequency in all 109 analyzed EEG electrodes of the three nights for each participant in the T/T and T/C group. The participant number is on the x-axis and the electrode number on the y-axis. Color-coding indicates the standard deviation of the selected peaks in Hz.

Because the distinct negative and positive peaks in the C3-RM power ratios indicate a shift in the spindle peak frequency rather than an overall increase in power spectral density, we next analyzed the periodic components of oscillatory spindle activity in the NREM sleep EEG. Intriguingly, the statistical analyses based on LMMs revealed a significant interaction between ‘exposure’ and ‘genotype’ for the center frequency of sleep spindle activity (χ^2^(Df) = 6.4(2), Pr(>χ^2^) = 0.04). Moreover, ‘exposure’ was a significant factor in the model for the center frequency (χ^2^(Df) = 7.6(2), Pr(>χ^2^) = 0.02) of sleep spindles.

### Spindle peak frequency specificity

As outlined in the methods section, we focused on the most pronounced peak within the spindle range. To verify the specificity of our selection procedure in case more than one peak was detected in that range, we computed the standard deviation of the center frequencies of the selected peaks in each electrode across the three nights of each participant as a measure of variability. Figure 3B illustrates the intra-participant spindle peak frequency variability in the T/T and the T/C allele carriers. In one participant in the T/T allele carriers’ group (Figure 3B, left panel, participant nr. 4) the variability in the selected center frequencies was higher in several electrodes (> 1.5 Hz in 32 out of 109 channels, with a total 109-channel mean ± standard deviation (std) of 0.85 ± 0.82 Hz). We arbitrarily chose this threshold because it may represent a minimum realistic frequency difference between slow and fast spindles within a participant. Upon visual inspection of the selected peaks in the highly variable channels, we confirmed the interchangeable selection of slow and fast spindle peaks (occurring due to overlap of the most prominent spindle peak with the lower peak selection threshold of 11 Hz). This participant was excluded from further analysis. Consequently, the mean standard deviation of the calculated standard deviations in the T/T carriers was 0.27 Hz, with a standard deviation of 0.08 Hz (n = 18), indicating the level of consistency of variability across channels and participants. Corresponding values for the T/C carriers were 0.20 ± 0.08 Hz (n = 15).

### High-density EEG analysis: Effects of 5G on center frequency of sleep spindles

Consequently, we performed topographical comparisons between the two exposure conditions and sham for both genotype groups and for all spindle peak variables extracted from the spectral parameterization. All-night data analysis did not reveal any significant differences between the conditions. The data depicted in Figure 4 refers to the first NREM sleep episode (combined stages N2 & N3, T/T carriers: n = 18, T/C carriers: n = 15). Statistical analysis of the sleep variables of the first sleep episode revealed an effect of ‘exposure’ on the duration of N1 sleep stage (χ^2^(Df) = 6.8(2), Pr(>χ^2^) = 0.03) but not for N2 and/or N3 (Supp. Table 6). Post-hoc t-tests did not indicate any significant differences between the groups and/or the experimental conditions. The center frequency of sleep spindles exhibited a widespread (i.e., 50/109 EEG channels) shift to higher frequencies in the T/C allele carriers after exposure to the 3.6 GHz field in a large cluster encompassing central, parietal, and occipital cortical areas. The percentage increase in the center frequency was 1.43 ± 6.5*10^-4^ %, corresponding to a mean shift in the frequency of the sleep spindle peak from 13.62 ± 0.1 Hz in the sham condition to 13.82 ± 0.1 Hz after the 3.6 GHz exposure (Figure 4). The acceleration in the center frequency after 3.6 GHz exposure in the T/C genotype was consistent and showed a large effect size (Cohen’s d mean ± std = 0.78 ± 0.18; Cohen’s d [min, max] = [0.28, 1.28]; Cohen’s d > 0.57 in 48 out of 50 channels). The topographical comparisons revealed no other effects of 5G RF-EMF exposure on the hd-EEG adjusted spindle power and bandwidth during the first NREM sleep episode (Figure 4).

**Figure 4.**
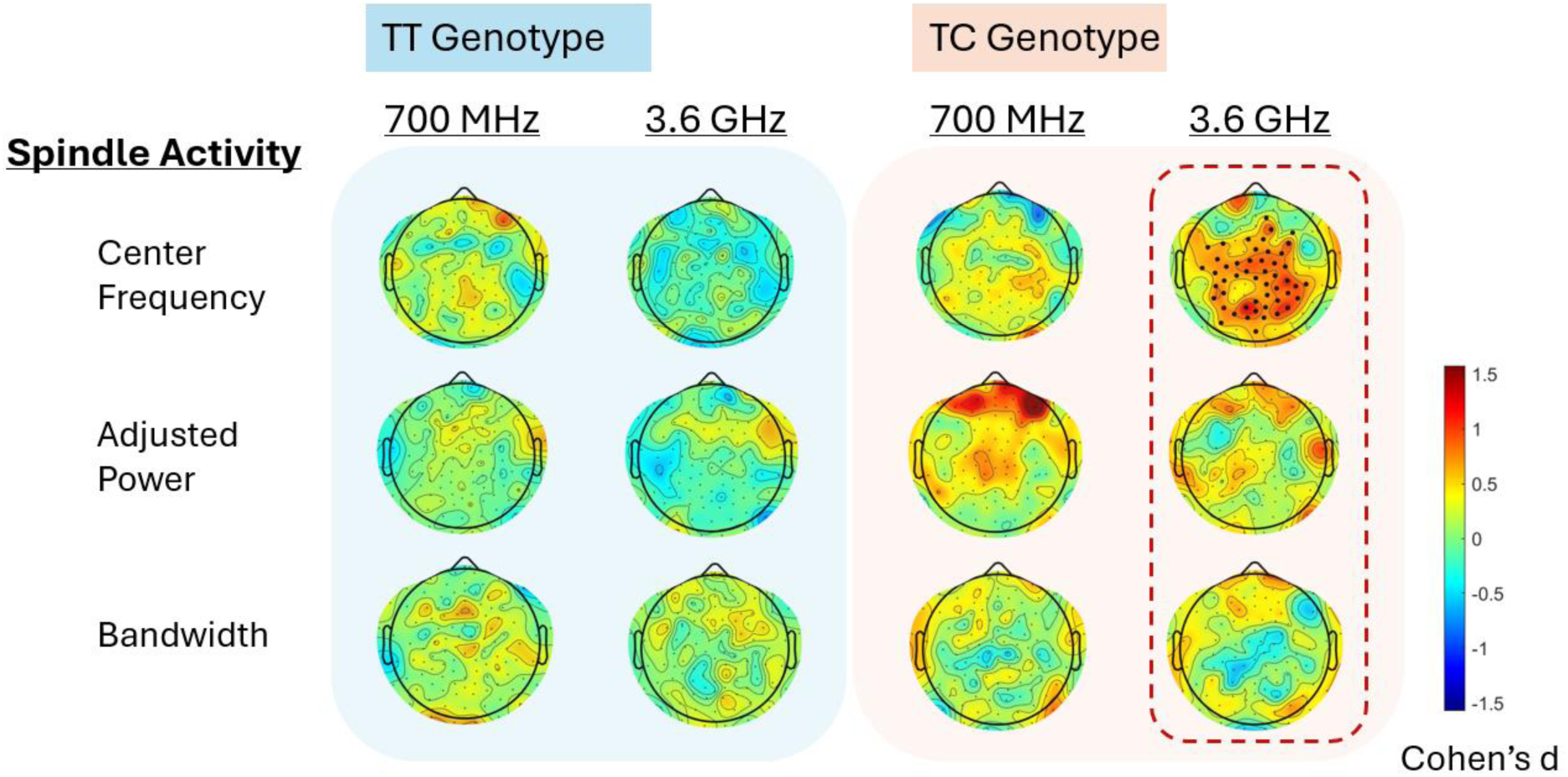
Topographical distribution of the effects of the two active fields (700 MHz and 3.6 GHz compared to sham) for both allelic variants (T/T: n=18 and T/C: n=15) for the extracted variables center frequency, adjusted power, and bandwidth in the first NREM sleep episode (combined stages N2 & N3). Cluster-based permutation statistics were applied, and significant cluster electrodes (p < 0.05) are indicated by black dots. The color bar indicates Cohen’s d effect sizes of the statistical comparison with sham exposure. Warm colors indicate an increase, and cold colors indicate a decrease compared to sham exposure.

## Discussion

We investigated whether two different radio-frequency EMFs of the newest 5G technology affect sleep and EEG sleep spindles in humans. In addition, we addressed for the first time the possibility of the contribution of an allelic variant (rs7304986) of the *CACNA1C* gene to the mediation of these effects. Benefitting from the recently developed FOOOF algorithm, we comprehensively characterized the 5G-induced changes in the spindle peak components in the NREM sleep EEG. Most importantly, we found a significant interaction between exposure and the genetic variant in the center frequency of sleep spindles. Specifically, we demonstrated a topographically widespread acceleration of spindle center frequency in the T/C carriers after exposure to the 3.6 GHz RF-EMF in comparison to sham.

Initially, we confirmed the spatial distribution of sleep spindle characteristics (Andrillon et al., 2011; Fernandez & Lüthi, 2020) following spindle peak spectral parameterization. More specifically, we found that spindle frequency is topographically organized with a sharp transition around the supplementary motor area between fast (∼13–16 Hz) centroparietal spindles and slow (∼11–13 Hz) frontal spindles, as it was previously reported with surface EEG, intracranial and unit recordings (Andrillon et al., 2011). It is important to note that with our peak selection process, we did not differentiate between slow and fast spindles in the same location/channel. The selection process identified the most pronounced peak within the sigma frequency range of the power spectrum as representation of the dominant spindle profile in the respective area and individual.

By leveraging the strengths of spectral parameterization, we demonstrated a widespread shift in the center frequency of sleep spindles towards faster oscillatory activity in T/C allele carriers after exposure to a 5G RF-EMF with a carrier frequency of 3.6 GHz. This effect was evident in central, parietal, and occipital cortical areas coinciding with areas that predominantly express faster spindles. A number of previous studies also reported enhanced EEG spectral power in the upper spindle range after exposure to RF-EMF of earlier generation (Huber et al., 2000, 2002; Schmid, Loughran, et al., 2012; Schmid, Murbach, et al., 2012). For example, controlled exposure to pulse-modulated RF-EMF signals of 2 W/Kg increased power between 13.75 – 15.25 Hz (Schmid, Murbach, et al., 2012) and 12.75 – 13.25 Hz (Schmid, Loughran, et al., 2012). However, these signals had a frequency of 900 MHz and their generation was based on a GSM frame structure as opposed to the latest 5G technology structure of the currently applied signals, which might introduce new sources of variability in the observed effects. Nevertheless, the earlier work was restricted to band-based power ratio analysis. It was not analyzed whether the previously observed changes in narrow-band spindle power also reflected a shift in the center frequency of spindles rather than an actual power increase, or both. Here we found in the power ratio analysis of the C3-RM channel ipsilateral to the exposure a pronounced peak at 14.75 Hz in the T/C carriers following exposure to the 3.6 GHz RF-EMF. The adjacent reduction in power is consistent with a shift in power within the spindle range rather than an increase in fast frequency spindle activity. A smaller shift was also observed after exposure to the 700 MHz signal. If present, the effects of the lower frequency field remain marginally detectable with the current methodology.

The discrepancy between the deeper penetration of the 700 MHz signal revealed by the simulated SAR distribution in the brain and the more pronounced effects on the EEG sleep spindles observed following exposure to the 3.6 GHz signal remains unclear. Notably, the pulse modulation, which has been identified as critical for the biological effects of RF-EMF (Huber et al., 2002), was identical at 12.5 Hz in both fields and the psSAR10gr was consistently set at 2 W/kg. The findings underscore the necessity for a comprehensive investigation into the complex characteristics of the new 5G signals. Furthermore, they may suggest that the dielectric and conductive properties of the tissues associated with the minor allele may not be adequately represented by the current simulation parameters. Alternatively, the observed effects may indicate a distinct mode of action that is unrelated to SAR distribution.

The generation of sleep spindles depends on reciprocal interactions between the thalamus and the cortex (Steriade et al., 1993; Steriade et al., 1993). The cortico-thalamic origin might explain the observed widespread, bilateral effect on the center frequency of sleep spindles following unilateral RF-EMF exposure. Bilateral effects were also reported in previous studies applying older-generation RF-EMF signals (Huber et al., 2000). Based on electrophysiological recordings, the frequency of sleep spindles is determined by the duration of the phasic hyperpolarization elicited by Ca^2+^-dependent rhythmic inhibitory postsynaptic potentials in thalamo-cortical relay neurons (Steriade et al., 1986; Steriade et al., 1993). As demonstrated in cats, a hyperpolarization lasting about 70 ms results in spindle frequencies of approximately 14-15 Hz, whereas a longer lasting hyperpolarization leads to lower frequencies (Steriade & Amzica, 1998). Furthermore, LTCCs are highly expressed in thalamocortical cells, interneurons, and reticular thalamic neurons in the rat brain, suggesting their important involvement in the regulation of calcium-dependent processes critical for thalamic functions (Budde et al., 1998). The shift in spindle center frequency observed in the T/C allele carriers after 3.6 GHz RF-EMF exposure may thus be linked to shorter hyperpolarization periods in the thalamic nuclei. However, because thalamic exposure, especially after 3.6 GHz exposure, is minimal, it appears rather unlikely that the interaction with the RF-EMFs occurred at the level of the thalamus. The simulated SAR distribution values indicate comparable levels of superficial absorption between the two fields. Taken together with the widespread bilateral acceleration of spindle activity, these findings suggest that the effects are likely occurring at the cortical level, subsequently enhanced by the reciprocal thalamocortical interactions involved in spindle generation.

Interestingly, Knoblauch and colleagues (Knoblauch et al., 2005) demonstrated the circadian regulation of spindle center frequency with a frequency reduction (from ∼13.85 to 13.7 Hz) coinciding with melatonin secretion. These circadian changes in EEG power spectra are not directly associated with the circadian variation in the duration of the sleep stages (Dijk et al., 1997). Thus, a circadian effect of RF-EMFs cannot be excluded (Ohayon et al., 2019). However, no circadian markers were assessed in the current study.

### Limitations

The study was conducted in a rather small homogenous sample of young, mainly female participants. Thus, the findings cannot be generalized. In addition, the study did not differentiate between slow and fast spindles, potentially oversimplifying the complexity of spindle activity. Further, the study assessed the effects of a single exposure. Long-term exposure and its cumulative effects were not explored, which could have different implications for sleep and underlying neural activity. Finally, the mechanistic underpinnings of allelic variant rs7304986 of *CACNA1C*, as well as the importance of other genetic and/or environmental factors in modulating individual RF-EMF effects on sleep and other physiological processes remain unknown.

## Conclusion and outlook

The repercussions of allelic variant rs7304986 for the functioning of Ca_v_1.2 channels are currently not known. Nevertheless, it is noteworthy that we replicated the observation in a previous genome-wide association study that T/C allele carriers subjectively report a longer time to fall asleep than T/T allele carriers (Byrne et al., 2013). Thus, this allelic variant appears to have a functional significance that should be further explored. In any case, our results provide first evidence that the LTCC Ca_v_1.2 plays a mechanistic role in the interaction between EMF and the human brain. This hypothesis can be further tested by studying the effects of RF-EMF on the sleep EEG after selective pharmacological modulation of these channels. The differential effects observed between the 700 MHz and 3.6 GHz exposures highlight the importance of considering signal characteristics and tissue properties in understanding RF-EMF interactions. Overall, our results provide new insights into the genetic and biophysical factors underlying RF-EMF effects on sleep, emphasizing the need for more targeted studies to elucidate these mechanisms.

## Supporting information

Sousouri et al_5G_Supplemental

## Data Availability

All data produced in the present study are securely stored in servers of the University of Zurich. Access and availability will be provided upon a material transfer agreement and after approval by the local ethics committee of the Canton of Zurich.

## Author contributions: CRediT

**Georgia Sousouri:** Data curation, Formal analysis, Investigation, Methodology, Validation, Visualization, Writing – original draft, Writing – review & editing. **Corinne Eicher:** Investigation, Data curation, Writing – review & editing. **Rachele Maria D’ Angelo:** Data curation, Writing – review & editing. **Marie Billecocq:** Investigation, Data curation, Writing – review & editing. **Thomas Fussinger:** Methodology, Software. **Mirjam Studler:** Investigation, Data curation, Writing – review & editing. **Myles Capstick:** Methodology, Validation, Software, Writing – review & editing. **Niels Kuster:** Resources, Software, Writing – review & editing. **Peter Achermann:** Validation, Writing – review & editing. **Reto Huber:** Resources, Validation, Writing – review & editing. **Hans-Peter Landolt:** Conceptualization, Funding acquisition, Resources, Supervision, Project administration, Investigation, Validation, Writing – review & editing.

## Declaration of generative AI and AI-assisted technologies in the writing process

During the preparation of this manuscript, the authors utilized ChatGPT-4 and Grammarly to assist in identifying and correcting spelling and grammatical errors in the text. Suggestions provided by these tools were carefully reviewed, and selected recommendations were incorporated. The authors affirm that they take full responsibility for the accuracy and content of the final manuscript.

## Acknowledgements

We would like to thank Fabio Carbone, MSc, Magdalena Kälin, MSc, and student interns for their dedicated contribution to the data collection and curation, Selina Schühle, MSc, for her help with data curation, and all study volunteers for their participation.

## Abbreviations

5G: Fifth-generation mobile telecommunication technology
RF: Radiofrequency
EMF: Electromagnetic fields
EEG: Electroencephalography
EOG: Electrooculography
EMG: Electromyography
NREM: Non-Rapid-Eye-Movement
LTCC: L-type Calcium Channels
CACNA1C: Gene encoding the α1C subunit of LTCC
FOOOF: Fitting Oscillations & One Over f
GSM: Global System for Mobile
hd: High density
SAR: Specific Absorption Rate
psSAR: Peak Spatial Specific Absorption Rate
ICNIRP: International Commission on Non-Ionizing Radiation Protection
TDD: Time Division Duplexing
FDD: Frequency Division Duplexing
OFDM: Orthogonal Frequency-Division Multiplexing
QPSK: Quadrature Phase Shift Keying
FIR: Finite Impulse Response
GUI: Graphical User Interface
SQM: Sleep Quality Marker
PSQI: Pittsburgh Sleep Quality Index
ESS: Epworth Sleepiness Scale
MCTQ: Munich Chronotype Questionnaire
EHS: Electro-Hypersensitivity
AASM: American Academy of Sleep Medicine
WASO: Wake After Sleep Onset
AIC: Akaike Information Criterion
CV: Critical Value
CI: Confidence Interval
SEM: Standard Error of the Mean

