## Supplementary material for "5G Radio-Frequency-Electromagnetic-Field Effects on the Human Sleep Electroencephalogram: A Randomized Controlled Study in *CACNA1C* Genotyped Healthy Volunteers": Sousouri et al_5G_Supplemental

#### Supplementary Figures

##### Exposure System

The sXh5G system is designed to expose a volunteer's head to either 700MHz or 3600MHz 5G modulated signals as defined in Table 1. The two carriers are not applied simultaneously. The general architecture of the system is depicted in Figure 1. A radiofrequency (RF) generator produces the modulated carrier that is then amplified and sent to the antenna matching the excitation signal frequency. The forward signal to the antenna is monitored using a power meter and a Schottky diode, as well as the reflected signal that is measured by a Schottky diode. This ensures the safety of the exposed participant.

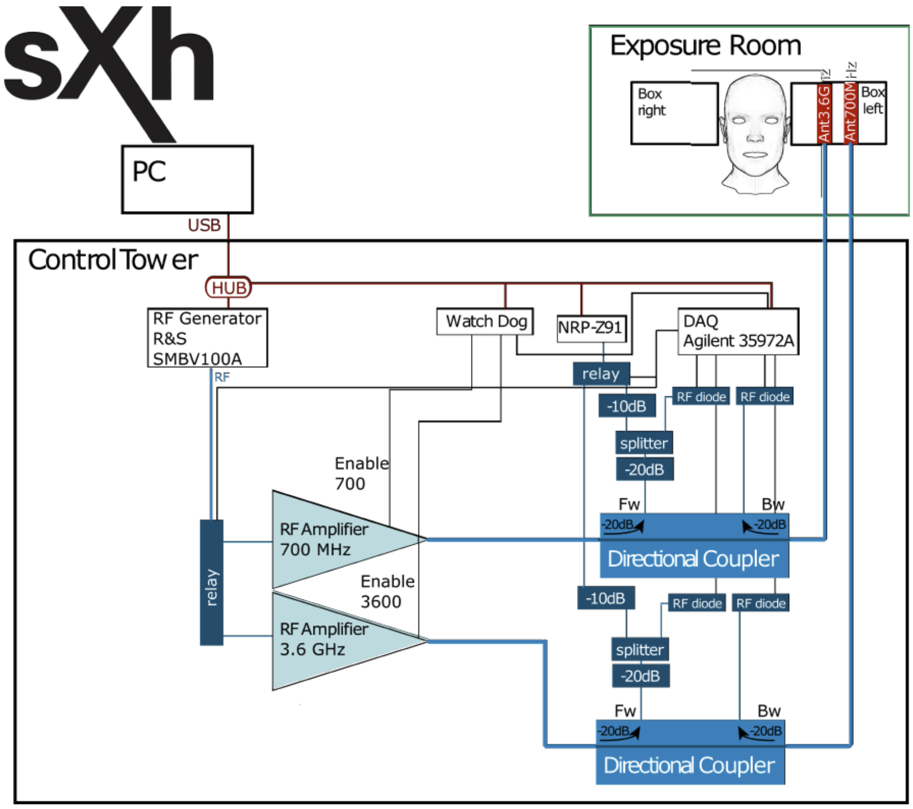

**Supplementary Figure 1. Architecture of the sXh5G exposure system.**

#### 5G Signal Characterization

The two signals with a carrier frequency of 700 MHz and 3.6 GHz were modulated to provide the same temporal variation in transmitted power. The 5G fields administered were uplink signals generated in the 5G frequency range. The lower frequency signal has a carrier frequency of 700 MHz, 20 MHz bandwidth, Frequency Division Duplexing/Orthogonal Frequency-Division Multiplexing (FDD/OFDM) with 24 resource blocks, 16 time slots, 60 kHz sub-carrier spacing and Quadrature Phase Shift Keying (QPSK) modulation, with an output power of 4.28 W to obtain 2 W/kg psSAR averaged over 10g. The higher frequency signal has a carrier frequency of 3.6 GHz, 100 MHz bandwidth, Time Division Duplexing (TDD)/QPSK OFDM with 135 resource blocks, 16 time slots, 60 kHz sub-carrier spacing and QPSK modulation, with an output power of 1.63 W to achieve the same psSAR. In the signals used all frames are identical with 16 time slots. Both signals have identical power control applied that introduces a repetitive low frequency amplitude modulation at 12.5 Hz on top of the pulse modulation due to the occupied time slots which have a dominant power modulation frequency of 200 Hz resulting in a 14.2 dB peak to average power ratio (PAPR).

The resulting amplitude variations in the time domain and low frequency envelope modulation components are illustrated in Supplementary Figures 2 and 3, respectively.

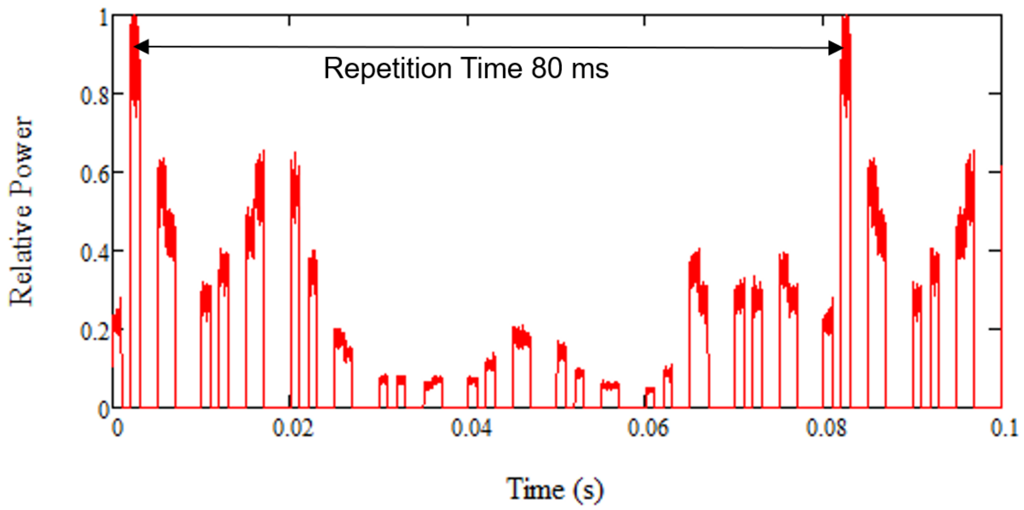

**Supplementary Figure 2. Repetitive amplitude variations due to slot occupancy and power control**

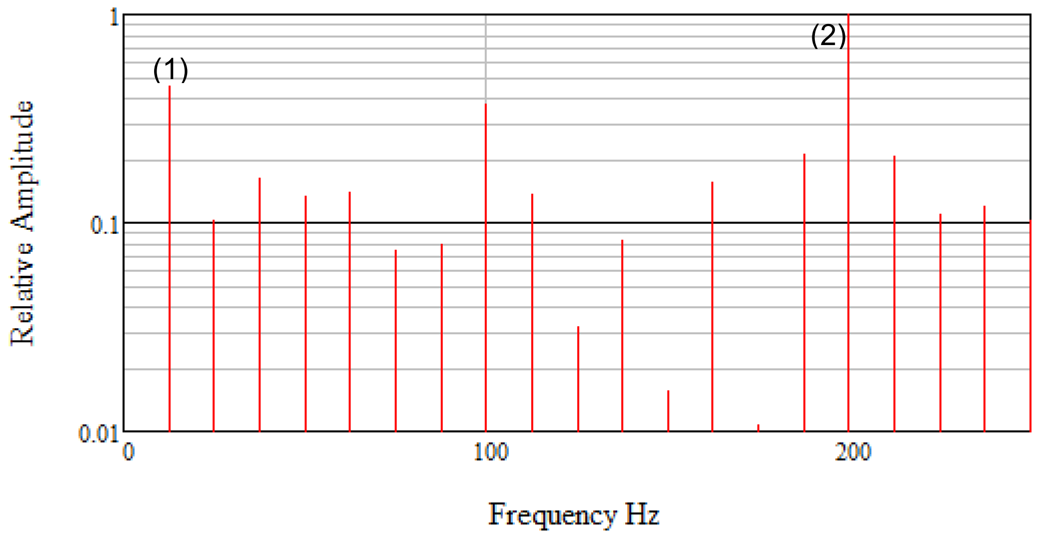

**Supplementary Figure 3. Low frequency envelope modulation components for both 700 MHz and 3.6 GHz signals.** (1) fundamental modulation component due to the applied power control, (2) pulse modulation due to the time slot length of 10 ms, other components are harmonically related to the power control.

#### Fitting Oscillations & One Over f (FOOOF) spectral parameterization

Supplementary Figure 3 (A-D) provides an illustrative description of the FOOOF algorithm for the parameterization of a neural power spectrum and the extraction of the aperiodic and periodic EEG components. In this example, three peaks are detected in the power spectrum, thus demonstrating the presence of three distinctive oscillatory components; one in the theta range (θ; 4-8 Hz), one in the alpha range (α; 8-11 Hz), and one in the sleep spindle, sigma frequency range (σ; 11-16 Hz).

#### FOOOF Gaussian Fitting Process

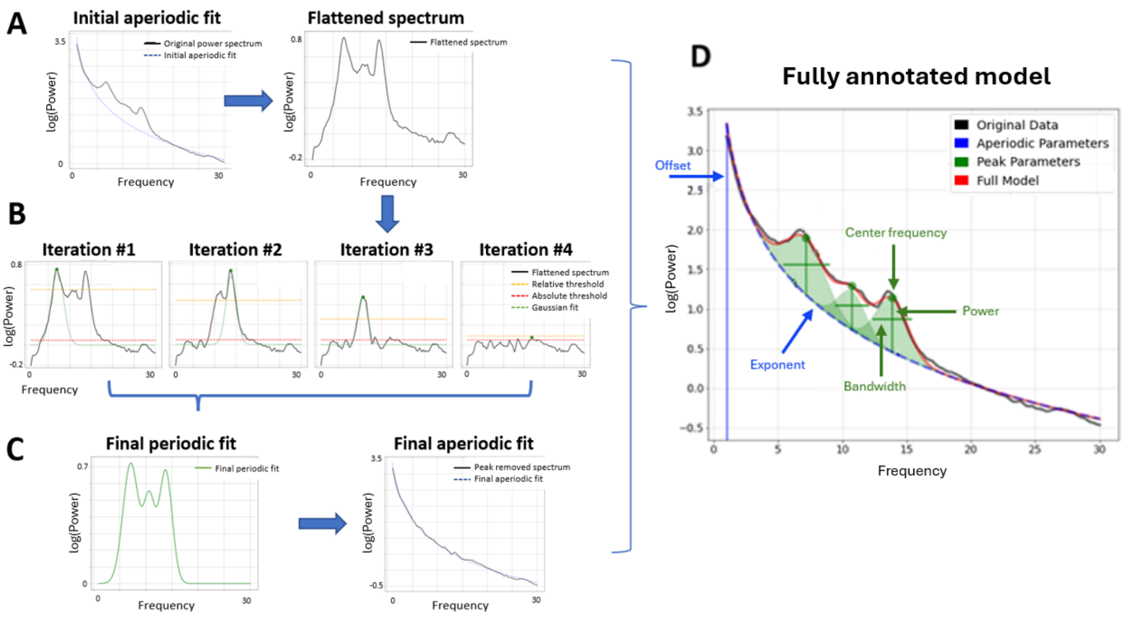

**Supplementary Figure 4. FOOOF Spectral Parameterization.** Application of the Fitting Oscillations & One Over f (FOOOF) algorithm to a sleep EEG power density spectrum. (A) Left: An initial fit of the aperiodic component is computed. Right: This aperiodic fit is subtracted from the power spectrum, leading to a flattened spectrum. (B) An iterative process identifies peaks in the flattened spectrum based on amplitude criteria and fits Gaussian distributions that best describe the morphology of each peak. The peak-search step halts when it reaches the noise floor, a relative threshold computed as two standard deviations of the flattened spectrum in units of the input data, re-calculated for each iteration (yellow line). This step is further controlled by setting a minimum absolute power threshold (red line). The power thresholds (relative or absolute) determine the minimum power beyond the noise floor that a peak must extend to be considered a putative oscillation. (C) Left: A final periodic fit is computed from all identified peak candidates. Right: The periodic peak fit is subtracted from the original power spectrum and a final fit of the aperiodic component is derived from the peak-removed power spectrum. (D) The full model is reconstructed from the combination of the aperiodic and peak fits. The aperiodic parameters are the offset (maximum log power at 1 Hz) of the whole spectrum and the exponent, describing the negative slope of the power spectrum in log-log space. The periodic parameters include the center frequency, the relative power (i.e., over the aperiodic component) and the bandwidth of each peak.

### Supplementary Tables

#### Simulations

SAR simulations were performed using SIM4LIFE (Zurichmedtech AG, Zurich Switzerland), the simulation parameters used in this study are summarized in Table 1. The grid is kept constant for all uncertainty and variability evaluations. The simulation is performed using anatomical heads chosen to match the age/sex variability described in the study proposal. As a next step, the anatomical models are replaced with the TwinSAM phantom using the same grid and simulation parameters to allow experimental verification of the simulated data. Free space simulation with the same grid allows evaluation of the antenna real-life power conversion efficiency.

**Supplementary Table 1.** Simulation Parameters.

| **Parameter** | | 700 MHz | 3600 MHz |
|---|---|---|---|
| General | number of periods:<br>auto-termination:<br>solver: | 90<br>None<br>aXware | 130<br>None<br>aXware |
| Excitation | type:<br>center frequency:<br>bandwidth: | edge source, Gaussian<br>700 MHz<br>600 MHz | edge source, Gaussian<br>3600 MHz<br>2000 MHz |
| Boundaries | type: | ABC, high | ABC, high |
| Grid | maximum step:<br>padding:<br>number of cells: | 0.5 mm<br>default, 93.69 mm in each direction<br>57MCells | 0.5 mm<br>default, 14.41 mm in each direction<br>49MCells |
| Voxeler | type:<br>maximum fraction: | Topological<br>21 | Topological<br>21 |

#### Combined SAR Variability

Table 2 shows the combined variability results for the whole brain based on 10g psSAR averages for five anatomical heads Duke (male, 34 years old), Ella (female, 26 years old), Fats (male, 37 years old), YoonSun (female, 26 years old) and the MIDA head

**Supplementary Table 2.** Combined variability

| | psSAR 10g | |
|---|---|---|
| Parameter | 700 MHz | 3.6 GHz |
| Head displacement (Frontal, Longitudinal, Antero-posterior, rotation, axial shift)* | 6.1% | 11.4% |
| Head model | 10.0% | 29.4% |
| **Combined variability (k=1) [%]** | 11.7% | 31.5% |

*Head distance relative to antenna (frontal axis) ±5mm, head height relative to antenna (longitudinal axis) ±10mm, head position relative to antenna (antero-posterior axis) ±10mm, head tilt relative to antenna all axes ±10 deg.

Table 3 shows the combined uncertainty for tissues of interest averaged over 0.125g which is approximately equal to a 5 mm cube.

**Supplementary Table 3.** Combined SAR variability including head displacement

| | Variability psSAR 0.125g | |
|---|---|---|
| Tissue(s) | 700 MHz | 3.6 GHz |
| Grey Matter | 14.0% | 39.9% |
| White Matter | 12.9% | 40.8% |
| White and Grey Matter | 13.6% | 38.6% |
| Whole Brain | 13.6% | 38.6% |
| Thalamus | 24.1% | 37.0% |

#### Exposure Uncertainty

Table 4 shows the combined uncertainty results for the whole brain. The experimentally related parameters relate to the experimental dosimetry performed to confirm the numerical dosimetry. The rest relates to the numerical dosimetry. Values are given for the whole brain peak spatial SAR over 10g (safety regulation related) and 0.125g (related more to the tissue volume distribution).

**Supplementary Table 4.** Combined uncertainty for the whole brain SAR

| | psSAR 10g | | psSAR 0.125g | |
|---|---|---|---|---|
| Parameter | 700 MHz | 3.6 GHz | 700 MHz | 3.6 GHz |
| Dielectric parameters ( $\sigma \pm 10\%$, $\varepsilon \pm 10\%$) | 0.3% | 8.9% | 4.5% | 4.3% |
| Signal bandwidth (20 MHz, 100 MHz) | 5.9% | 2.3% | 5.7% | 5.6% |
| **Experiment related (k=1) [%]** | 5.9% | 9.2% | 7.3% | 7.0% |
| Simulation time (80 -> 130, 160 -> 260 periods) | 0.0% | 0.2% | 0.0% | 0.2% |
| Grid resolution (1 mm -> 0.5 mm) | 0.2% | 3.1% | 0.5% | 2.3% |
| Padding size (x2) | 0.0% | 0.0% | 0.0% | 0.0% |
| **Simulation related (k=1) [%]** | 0.2% | 3.1% | 0.5% | 2.3% |
| **Combined uncertainty (k=1) [%]** | 5.9% | 9.7% | 7.3% | 7.4% |

#### Sleep variables

**Supplementary Table 5:** Wald Chi-squared test statistics for the factor ‘night’ in all-night sleep variables

| | $\chi^2$ (Df) | Pr(>$\chi^2$) |
|---|---|---|
| Total sleep time (min) | 18.3 (2) | < 0.01 |
| Sleep efficiency (%) | 18.7 (2) | < 0.01 |
| Sleep latency (min) | 12.4 (2) | < 0.01 |
| N1 sleep (min) | - | - |

| | | |
|---|---|---|
| N2 sleep (min) | - | - |
| N3 sleep (min) | - | - |
| N2 & N3 sleep (min) | 6.7 (2) | 0.03 |
| REM sleep (min) | 8.6 (2) | 0.01 |
| REM sleep latency (min) | 6.3 (2) | 0.04 |
| WASO (min) | 19.1 (2) | < 0.01 |

*Total sleep time = total amount of time spent asleep; Sleep efficiency = (total sleep time/time in bed) * 100; Sleep latency = time between lights-off and first occurrence of non-rapid-eye-movement (NREM) sleep stage N2; N1, N2, N3 = NREM sleep stages; N2 & N3 = combined NREM sleep stages N2 & N3; REM sleep = rapid-eye-movement sleep; REM sleep latency = time between sleep onset and first occurrence of REM sleep; WASO = wakefulness after sleep onset;* $\chi^2$ *= Wald Chi-squared test statistics for the factor 'night'; Df = degrees of freedom; Pr(>*$\chi^2$*) = p-value for the factor 'night'. '-' means that the factor 'night' was not included in the final model.*

**Supplementary Table 6:** Sleep parameters of the first sleep episode for the three experimental conditions of both genotypes

| *CACNA1C* genotype | **T/T allele carriers (n = 19)** | | | **T/C allele carriers (n = 15)** | | | |
|---|---|---|---|---|---|---|---|
| Exposure condition | **S0** | **E7** | **E3** | **S0** | **E7** | **E3** | $p_{Exposure}$ |
| N1 sleep (min) | 4.3 ± 2.1 | 4.7 ± 2.9 | 5.2 ± 2.8 | 3.7 ± 1.9 | 4.5 ± 2.5 | 5.6 ± 6.8 | 0.03 |
| N2 sleep (min) | 40.7 ± 18.4 | 37.1 ± 18.4 | 35.5 ± 18.3 | 38.1 ± 19.2 | 43.1 ± 28.7 | 36.6 ± 27.4 | - |
| N3 sleep (min) | 47.0 ± 20.5 | 42.7 ± 18.0 | 49.6 ± 17.7 | 49.4 ± 14.8 | 41.2 ± 10.9 | 41.0 ± 8.5 | - |
| N2 & N3 sleep (min) | 87.7 ± 29.2 | 79.8 ± 27.9 | 85.1 ± 25.6 | 87.4 ± 29.3 | 84.3 ± 28.1 | 77.5 ± 28.7 | - |
| REM sleep (min) | 9.5 ± 8.4 | 10.1 ± 8.7 | 9.1 ± 10.9 | 8.3 ± 6.4 | 7.1 ± 5.0 | 7.2 ± 9.7 | - |
| WASO (min) | 1.3 ± 2.7 | 0.8 ± 1.5 | 1.2 ± 1.3 | 2.0 ± 3.3 | 2.1 ±2.8 | 4.7 ±17.1 | - |

*Values represent means ± standard deviations. CACNA1C genotype: T/T and T/C allele carriers of gene variant rs7304986. Exposure conditions: S0 = sham exposure; E7 = 700 MHz carrier frequency, 20 MHz bandwidth; E3 = 3600 MHz carrier frequency, 100 MHz bandwidth. N1, N2, N3 = NREM sleep stages; N2 & N3 = combined NREM sleep stages N2 & N3; REM sleep = rapid-eye-movement sleep; WASO = wakefulness after sleep onset.* $p_{Exposure}$ *= Factor 'exposure' of linear mixed model (Wald Chi-squared test statistics). '-' means that the factor was not included in the model.*
